# Temporal changes in mental response and prevention patterns, and their impact from uncertainty stress during the transition in China from the COVID-19 epidemic to sporadic infection

**DOI:** 10.1101/2021.06.18.21259062

**Authors:** Sihui Peng, Xiaozhao Yousef Yang, Tingzhong Yang, Weifang Zhang, Randall R Cottrell

## Abstract

**Objective:** This prospective observational study examined changing trends of mental responses and prevention patterns, and their impact from uncertainty stress during the transition in China from the COVID-19 epidemic to sporadic infection.

**Setting:** A prospective longitudinal observation design was utilized in this study.

**Participants:** We recruited participants for an online panel survey from chat groups on Chinese social media platforms.

**Data collection:** There were 7 waves of interviews. Data were obtained by an online survey. A special administrative WeChat group was established to manage the follow-up data collection.

**Measures:** Several mental responses and prevention patterns were each measured by single questionnaire items. Uncertainty stress was measured by 5-point scale. An irrational beliefs about prevention variable was comprised 5 common misconceptions, which manifested during the COVID-19 epidemic in China.

**Analysis:** Sixty-two participants completed all observation points and were included in the study. The Mann–Kendall Test was used to assess changing trends across the seven observation points. The nonparametric linear mixed effects model was used to examine the association between uncertainty stress and mental and behavioral responses.

**Results:** The mean uncertainty stress did not change significantly over the observation period (T:-0.911, P>0.05). This trend was also true for perceived risk (T: -0.141, P>0.05), perceived severity (T: 1.010, P>0.05), self-efficacy for prevention (T: 0.129, P>0.05), and prevention behavior (T: 0.728, P>0.05). There was a statistically significant downwards trend in irrational beliefs about prevention (T: -4.993, p < 0.01), sleep (T: -2.499, p < 0.05), emotions (T: -5.650, p < 0.01), and lifestyle (T:-5.978, p < 0.01). The results showed that uncertainty stress was positively associated with irrational beliefs (β: 0.16298, p<0.01). The more uncertainty stress, the worse was their sleep (β: 0.02070, p<0.05), emotions (β: 0.03462, p<0.01), and lifestyle(β: 0.02056, p<0.05). High levels of uncertainty stress was negatively associated with self-efficacy for prevention and prevention behavior, βvalue was =-1.33210 (p<0.01) and -0.82742 (p<0.01), respectively.

**Conclusion:** As the COVID-19 virus spreads around the globe, it is currently in epidemic status in some countries, in sporadic status in another countries, and it will eventually transition to a sporadic infection status. This study provides new information on changing trends of mental responses and prevention patterns from the COVID-19 epidemic as the transition to a sporadic infection period takes place. These results may have important policy and disease prevention in post-epidemic times.

## Introduction

The COVID-19 pandemic is a public health emergency that has caused unprecedented disruption to the way most people live and work. The disease induced strong mental and behavioral response that may lead to significant mental and health disorders among large segments of the population. According to the Stimulus, Cognition and Response (SCR) model, various stimuli (S) affect the internal states of people through cognition (C), which in turn elicits mental and behavioral responses (R) (Cottrell and McKenzie, 2010, Yang, 2018). COVID-19 is a strong stimulus that plausibly induces people to perceive high risk of infection with potentially severe health consequences(Organization, 2020). Many studies found that the potential to contract highly lethal disease, such as COVID-19, can overwhelm people emotionally and physically, and induce strong mental and behavioral responses (Greenberg et al., 2020, Cullen et al., 2020, Li et al., 2020, LV S and Yang, 2008). Given the importance of human mental and behavioral functioning to disease control, it is crucial to study these factors during and after an outbreak (Bish and Michie, 2010, Wise et al., 2020, Yang et al., 2021). Currently, COVID-19 is still in epidemic status in many countries around the world, in sporadic status in other countries, and it will eventually transition to a sporadic infection status. It is of special significance to understand the impact of time on mental and behavioral reactions to COVID-19 as this may have implications on policy and possible behavioral interventions. However, most studies of COVID-19 were cross-sectional, and few previous empirical studies utilized longitudinal observation(Cerami et al., 2020, Yang et al., 2021). A prior prospective observational study examined changing trends in mental and behavioral responses during the epidemic phase of COVID infection in China. At the time of this study, COVID-19 was causing sporadic infections in China. It was not known how a person’s mental and behavioral responses, included prevention beliefs and behavior might change as the incidence of the disease decreased. The main purpose of this research is to evaluate changing trends in mental responses and prevention patterns during this period in China. This is urgently needed to inform disease prevention specialists, especially those who work with the most vulnerable populations.

Uncertainty occurs when an event or a situation causes ambiguity, inconsistency, or unpredictability (Mast, 1995, Yang, 2018). COVID-19 is a new virus. Much information is unknown or imperfect, and there is much ambiguity surrounding the disease. An outstanding characteristic of the COVID-19 epidemic is uncertainty. A host of evidence supports the assertion that uncertainty constitutes a powerful stressor(Dar et al., 2017, Greco and Roger, 2003, Yang, 2018). Some authors have noted that uncertainty stress has had a negative impact on COVID-19 epidemic control and has created challenges for disease prevention(Brown, 2020, Petterson et al., 2020). One recent study found that uncertainty stress was an independent stimulus in the SCR theoretical framework (Peng et al., 2021). Uncertainty stress has been at persistently high levels throughout the COVID-19 epidemic, and it has been positively associated with disease fear, and negatively associated with self-efficacy and prevention behaviors. Currently, it is not known if these higher levels of uncertainty stress will be maintained as COVID-19 enters into a more sporadic infection period. Will it still contribute to mental and behavioral responses, especially, prevention patterns? This study will seek to answer these questions.

The study may yield information to assist in formulating evidence-based public policy decisions, directing prevention efforts and implementing health education initiatives aimed at mental and behavioral problems in post-epidemic time. The information may also be of significance for future epidemics and pandemics that may occur.

## Methods

### Study design

A prospective longitudinal observation study was designed to examine temporal trends and changes in mental and behavioral responses as COVID-19 transitioned from epidemic status to a period of more sporadic infections, and their associations with uncertainty stress in China.

### Participants

Participants were recruited via a survey advertisement in the social media groups WeChat and Douban, two of the most popular social media platforms in China. Inclusion criteria were membership in a common community; being between 20-60 years of age; having access to a Smartphone; knowing the Chinese language; and willing to participate in the panel study and provide follow-up information at seven scheduled observation points. Participants were excluded if they refused to provide this information or had a medical condition that could limit or preclude their participation. Within the registration system, potential participants were screened to ascertain eligibility. Upon consent, participants received an electronic questionnaire and instructions on how to proceed. After reading the instructions, they were asked to provide an e-consent by tapping the “Confirmation and Authorization” button and then directed to the questionnaire. A special administrative WeChat group was established to manage the follow-up data collection, using a unique QR code for each respondent. The QR code was the vehicle, not only for identifying unique participants, but prohibiting non-participants from taking the survey. After scanning the QR code, survey participants could enter the investigation group without further preconditions.

This panel study includes seven waves of data collected from COVID-19 epidemic status to the sporadic infection period. The data collection time interval of each wave was 7 days. Data were collected at two different times; early in the year February and March 2020 and later in the year, December 2020. The actual dates of the data collection were as follows: wave 1(5/Feb/2020), wave 2(12/Feb/2020), 3(19/Feb/2020), wave 4 (26/Feb/2020), and wave 5(4/March/2020) in the first period. The corresponding number of reported new confirmed patients in this period in China respectively numbered 3,887, 2,015, 394, 433, and 133(National Health Commission of People’s Republic of China, 2021). The second period of data collection was: wave 6 (23/December, 2020) and wave 7(30/December, 2020). The corresponding number of reported new confirmed patients in this period respectively numbered 17 and 25 (China, 2020).

### Data Collection

Data was obtained by an online survey, to ascertain mental and behavioral responses to COVID at that time. An online survey was implemented on *Wenjuanxing* (www.wjx.cn), a survey service website similar to Qualtrics or Survey monkey, but tailored to Chinese users. Each wave of the survey had a dedicated electronic questionnaire access link. The online questionnaire link was posted to the respondent group, centrally managed in a WeChat group, and accessible every Wednesday from 10:30 am to 4:30 pm. Data were collected from 9.00-11.00 am every Monday. Data collectors and facilitators were third-year doctoral students enrolled in a university public health program. All responses were anonymous. The questionnaire took approximately 10 minutes to complete, and the same survey protocol was used for each wave of the survey to assure homogeneity of data administration and collection. As appropriate, a token of appreciation, 30 RMB (approximately$5.00 US dollars) was given to those participants who completed all 7 questionnaires.

### Measurement

In this study, basic individual demographic characteristics were recorded as age, sex, ethnicity, education level, marital status and occupation.

Mental and behavioral variables about prevention in this study included several factors, as described below:

*Perceived risk and severity, emotional, sleep, and lifestyle problems* are common mental responses. (1) Perceived risk was measured by a question “Do you always feel that you may be infected? Responses were on a 5-point Likert-type scale ranging from “strongly disagree” to “strongly agree.” Perceived severity was measured by the question “If you were infected with COVID-19 it would be a serious misfortune.” Responses were again on a 5-point Likert-type scale ranging from “strongly disagree” to “strongly agree”. (2) Emotional status, sleep and lifestyle status were measured by three questions, that asked, “how is your current (emotional, sleep, lifestyle) status as compared to before COVID-19 epidemic?” Respondents were to select one of three options: “same as before” “less stable than before” or “much less stable than before.”

#### Uncertainty stress

COVID-19 is an unfamiliar, infectious disease associated with high mortality rates. Uncertainty stress is the most common and strongest among different types of mental stresses (Peng, et al, 2021). Uncertainty stress was measured utilizing a scale designed by Yang and colleagues (Yang, et al, 2019), which has demonstrated acceptable validity, and has since been used extensively in Chinese research (Yang, et al, 2019; Wu, et al, 2020). It covered 4 items; current life uncertainty (“life is unstable and cannot be controlled”), social change uncertainty (“uncertain about what will happen in the future”), goals uncertainty (“uncertain about how to achieve goals”) and social values uncertainty (“cannot follow social values”). Respondents rated these items on a 5-point scale from feel no stress (0), a little stress (1), some stress (2), considerable stress (3), and very strong stress (4). A total stress score was obtained by adding up the responses to the individual questions. The higher the total score, the greater the perceived level of uncertainty stress (Wu et al., 2020, Yang et al., 2019).

#### Irrational beliefs

Irrational beliefs are manifested from people’s mental response under environmental pressure, especially high pressure (Yang, 2018). Some studies found that during COVID-19 epidemic, people showed more irrational beliefs than would be typical (Teovanović et al., 2021, Oliver and Wood, 2014). Unusual environmental threats cause higher stress levels in people which is associated with impaired reality and strange thinking patterns (McNAUGHTON et al., 1995, Amutio and Smith, 2008). This may lead to irrational beliefs about prevention with increased levels of perceived risk and severity of COVID-19 (Teovanović et al., 2021, Sironi et al., 2020). We refer to irrational beliefs as an umbrella term that covers beliefs which lack a solid evidence base or defy principles of normative rationality (Dorfan and Woody, 2011, Rabalais, 2015). These beliefs are self-defeating, unconditional, in conflict with reality, and unlikely to find empirical support. This study addressed irrational beliefs about effective COVID-19 prevention measures in China that were not founded upon reality and science. They included, (1) smokers are not susceptible to COVID-19, (1) consuming alcohol can prevent the spread of the virus, (3) people should avoid people from Hubei province, where COVID-19 first manifested in China so as to prevent contraction of the disease, (4) employees from Hubei should be dismissed to prevent the spread of the novel virus, and (5) people who move away from an affected area should be deported back to their place of origin. Items were rated on a 5-point Likert-type scale, ranging from 1 (strongly disagree) to 5 (strongly agree). Item scores were summed to attain a total score for belief in COVID-19 prevention myths. The analysis showed that the Cronbach’s α coefficient of this scale was 0.73 suggesting acceptable reliability. One factor was extracted at each wave, accounting for 66% of the variance. Factor loading values among the five items was 0.76, 0.75. 0.65, 0.59, and 0.49, indicating the scale has good construct validity.

#### Self-efficacy for prevention and prevention behavior

Self efficacy is commonly defined as the belief in one’s capabilities to achieve a goal or an outcome. Self-efficacy for preventing COVID-19 infection was measured by asking, ‘Do you think that you can avoid the disease through your current prevention behaviors?’ Responses were provided on a 5-point Likert type scale ranging from ‘no confidence I can avoid the disease’ to ‘much confidence I can avoid the disease’. Prevention behaviors against COVID-19 infection was measured by a question, “I feel my precautions against infection by Covid-19 were”, with options on a 5-point scale ranging from “very weak” to “very strong”.

It should be noted that some of the above variables used a single questionnaire item to measure the variable. This has been used in previous literature, and especially in other COVID-19 studies(De Zwart et al., 2009, Tomczyk et al., 2020, Yildirim and Güler, 2020). There is also anecdotal evidence to suggest the validity of using one question measures of these variables. Previous studies have shown that those responding positively to these questions were more likely to take precautionary action(De Zwart et al., 2009, Tomczyk et al., 2020, Yildirim and Güler, 2020).

### Data analysis

All data were entered into a database using Microsoft Excel. They were then imported into SAS (9.4version) for the statistical analysis. Across survey waves mean scores were calculated for the various variables at different observation points. As most of the variables included in this study were not normally distributed we used nonparametric testing methods to examine changing trends in mental and behavioral responses, and their associations. The Mann–Kendall Test was used to assess changing trends across the seven observation points (Kendall, 1948). The nonparametric linear mixed effects model was used to examine the association between uncertainty stress and several mental and behavioral response variables (Trong et al., 2020). Regression parameters in fixed effects were estimated using the Theil-Sen test (Fernandes and Leblanc, 2005, Q, 2008).

## Results

One hundred-and-fifty participants were recruited at baseline. The baseline was linkable to five intermediate and a final observation point, with 102 participants completing the survey during the COVID-19 epidemic period, 62 completing the survey during the COVID-19 sporadic infection period. Sixty-two participants completed both the baseline and final observation points and were included in the analysis. Of the study sample, 43.0% were female and 91.9% were Han Chinese. Those less than 30 years old comprised 41.9% of the sample and 39.9% were more than 50 years of age. Forty were female and a half percent was never married. High school or junior college graduates made up 54.7% of participants while 46.8% had completed an undergraduate degree or higher. As to occupation, 24.2% were managers, 19.4% were professionals, 21.0% were commercial or service workers, 11.3 were operators, and 24.2% were retired. (Table 1).

**Table 1.** Characteristics of sample

| Group | N (62) | % |
| --- | --- | --- |
| Sex |  |  |
| Male | 19 | 30.6 |
| Female | 43 | 69.4 |
| Age (years) |  |  |
| <30 | 26 | 41.9 |
| 30-49 | 15 | 24.2 |
| 50+ | 21 | 33.9 |
| Education |  |  |
| High school or junior college | 32 | 51.6 |
| College and more | 30 | 48.4 |
| Ethnicity |  |  |
| Han | 57 | 91.9 |
| Minority | 5 | 8.1 |
| Marital status |  |  |
| Never married | 28 | 45.2 |
| Married | 29 | 46.8 |
| Divorce or widowed | 5 | 8.0 |
| Occupation |  |  |
| Manager | 15 | 24.2 |
| Professional | 12 | 19.4 |
| Commercial or service | 13 | 21.0 |
| Operator | 7 | 11.3 |
| Retired | 15 | 24.2 |

**Table 2.**
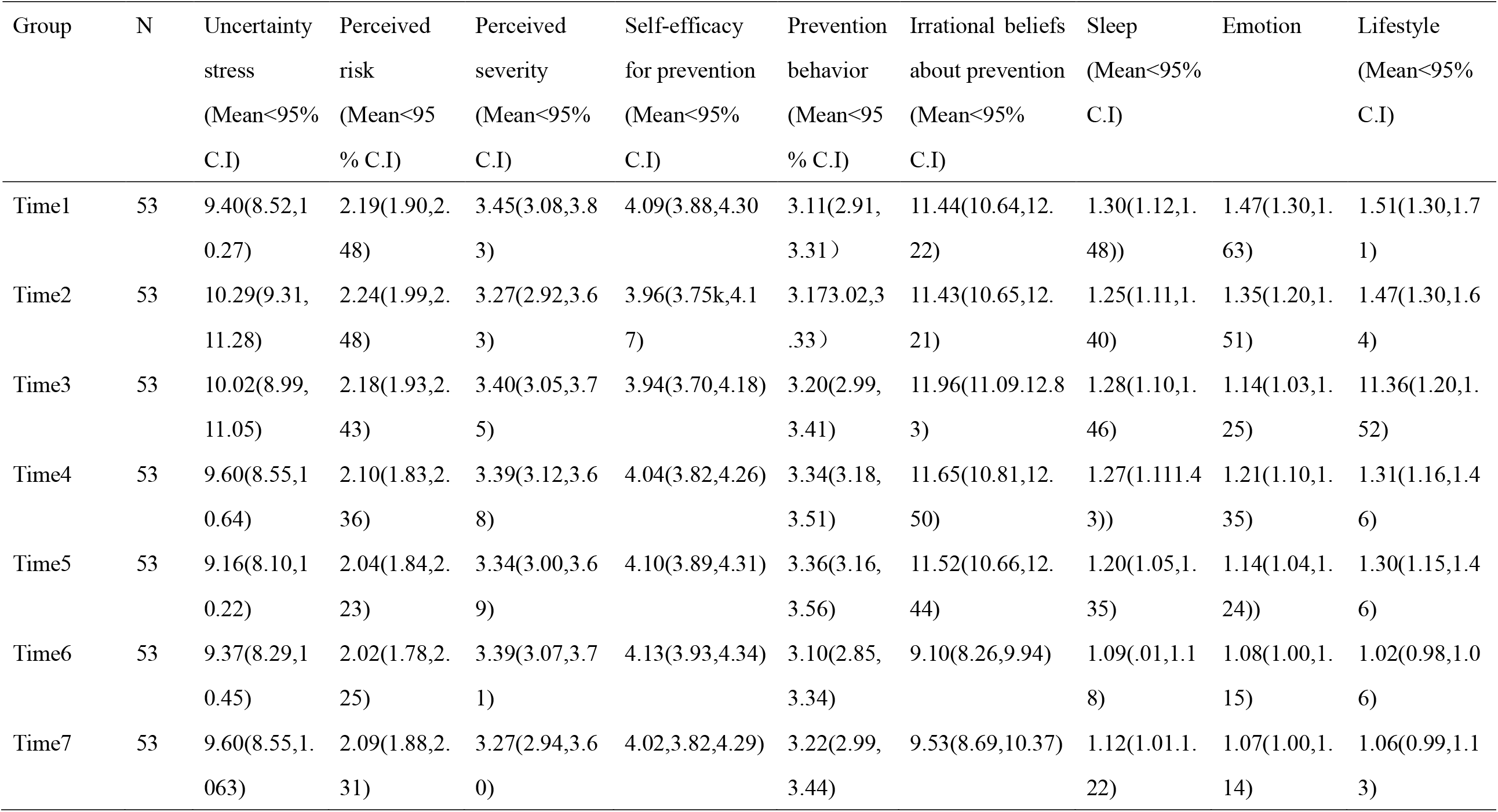
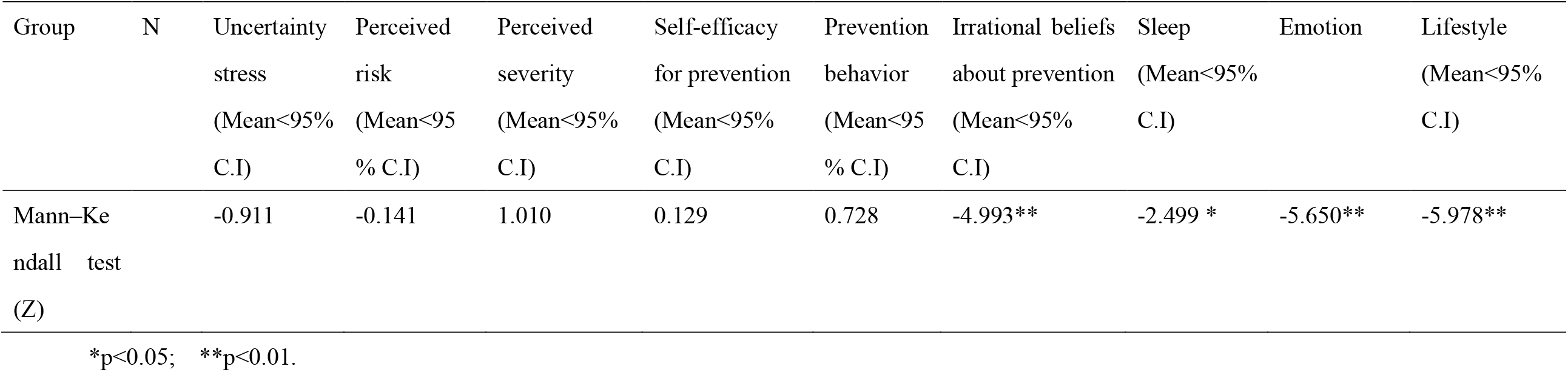
Means and their change trend across time in metal and behavioral response variables

The mean uncertainty stress score at baseline was 9.40 and it was 9.60 at the end of the study, which did not change significantly over the observation period (T:-0.911, P>0.05). This trend was also same for perceived risk (T: -0.141, P>0.05), perceived severity (T: 1.010, P>0.05), self-efficacy for prevention (T: 0.129, P>0.05), and Prevention behavior (T: 0.728, P>0.05). There was a statistically significant downwards trend in irrational beliefs about prevention (T: -4.993, p < 0.01), sleep (T: -2.499, p < 0.05), emotion (T: -5.650, p < 0.01), and lifestyle (T: -5.978, p < 0.01).

Table 3 indicates that uncertainty stress was positively associated with irrational beliefs(β: 0.16298, p<0.01). The more uncertainty stress, the worse was their sleep(β: 0.02070, p<0.05), emotion(β: 0.03462, p<0.01), and lifestyle(β: 0.02056, p<0.05). Higher uncertainty stress levels were negatively associated with self-efficacy for prevention and prevention behavior, βvalue was =-1.33210 (p<0.01) and -0.82742 (p<0.01), respectively.

**Table3.** Association of Uncertainty Stress and irrational beliefs about prevention, self-efficacy for prevention, prevention behavior, sleep, emotion, and lifestyle

| Group | Estimate($\beta$) | Standard Error (S.E) | P |
| --- | --- | --- | --- |
| Irrational beliefs about prevention | 0.16298 | 0.06080 | 0.007781 |
| Self-efficacy for prevention | -1.33210 | 0.23269 | <0.0001 |
| Prevention behavior | -0.82742 | 0.25419 | 0.0001 |
| Sleep | 0.02070 | 0.00815 | 0.0115 |
| Emotion | 0.03462 | 0.00644 | <0.0001 |
| Lifestyle | 0.02056 | 0.00821 | 0.0127 |

## Discussion

Addressing a gap in the literature, this study found temporal trends in mental responses and prevention patterns from the COVID-19 epidemic to the sporadic infection period. There was a statistically significant downwards trend in sleep, emotion, and lifestyle problems. This phenomenon may be explained by SCR theory. COVID-19 is a strong stimulus that plausibly induces mental and behavioral responses during an epidemic. Studies have reported on high mental and behavioral responses during the COVID-19 epidemic (Cullen et al., 2020, Greenberg et al., 2020, Li et al., 2020, LV S and Yang, 2008). It might be expected that some mental and behavioral responses, sleep, emotion, and lifestyle problems, would be reduced. As COVID transitioned from the epidemic phase to the sporadic infection phase. Further, irrational beliefs were common during the COVID-19 epidemic. Irrational beliefs are self-defeating, unconditional, in conflict with reality, and are not supported by empirical data. They may, in fact, exacerbate the epidemic by negatively impacting preventive behaviours (Dorfan and Woody, 2011, Teovanović et al., 2021). Irrational beliefs appeared very quickly when COVID-19 infections began, and dropped rapidly as the epidemic declined and the sporadic infection phase was apparent.

Perceived risk and severity did not show significantly downward trends in this transition period. This indicates that people still maintained a high alert status for COVID-19 infection. Self-efficacy is commonly defined as the belief in one’s capabilities to achieve a goal or an outcome. This construct also remained relatively stable during the study period. This study found no significant temporal changes in preventive behavior. Overall these data seem to indicate that people were still concerned about COVID-19 and their ability to prevent it even as the diseased moved from an epidemic status to a sporadic infection status.

This study did find that the high levels of uncertainty stress persisted from the COVID-19 epidemic phase to the sporadic infection period. Uncertainty occurs when an event or a situation causes ambiguity, inconsistency, or unpredictability. The COVID-19 outbreak created a great deal of uncertainty. High uncertainty stress would be expected and was documented during the COVID-19 epidemic phase(Peng et al., 2021). It might be expected that uncertainty stress would be reduced as the epidemic progressed and would especially be reduced in sporadic infection period when infection risk is reduced. This was not the case. High levels of uncertainty stress in the sporadic infection phase indicate that there are other challenges besides COVID-19 infection numbers in this process.

Although the disease itself declined, the epidemic has had a strong negative impact on economic activity and production. This has created much uncertainty in employment, work and life. Moreover, this study found that uncertainty stress was positively associated with “instantaneous” responses, and was negatively associated with “stable” responses. This indicates uncertainty stress remains a large challenge to prevention in the COVID-19 sporadic infection period. This would imply that COVID-19 interventions should focus attention on decreasing uncertainty stress and on improving production and economic recovery.

There are several limitations to this study. First, our sample size was small. Nevertheless, this is a prospective longitudinal panel study, and the variables included were repeatedly measured for each participant. Statistical power for the tests used in this study was high, so the sample size was large enough to make appropriate inferences. Second, sample attrition may introduce “cluster” bias because many longitudinal studies likely over-represent some of these characteristics, such as high educational attainment. A more sophisticated design and representative sample would be necessary to resolve this problem. Third, high non-response rate (39.2%) occurred in the second stage follow up. This could, in part, be due to the long period of time from the first stage to the second stage. But, demographic characteristic comparisons between the first and second stage samples(Peng et al., 2021). Further studies are necessary to reduce the bias.

As the virus spreads around the globe, COVID-19 is still in epidemic status in many countries, while it moves from epidemic to sporadic infection status in other countries. This study provides new information on changing trends of mental responses and prevention patterns from the COVID-19 epidemic phase to the sporadic infection period. The study may have important implications for policy and disease prevention for many countries of the world.

## Data Availability

The study protocol and additional data can be shared on request from the corresponding authors.

## Acknowledgement

This study was supported, in part, by the National Nature Science Foundation of China (71490733) and Natural Science Foundation of Guangdong Province of China (Grant No. 2018A030307002).

## Notes

### Competing Interest Statement

The authors have declared no competing interest.

### Author Declarations

This study was approved by the Ethics Committee at the Medical Center, Zhejiang University (2014: 1-017)

